# Challenging the guidelines: Longitudinal Trends in Left Ventricular Diameter and Function in Severe Aortic Regurgitation

**DOI:** 10.64898/2026.04.09.26350549

**Authors:** Shmuel Schwartzenberg, Amit Berkovitz, Tsahi T. Lerman, Tamir Bental, Mordehay Vaturi, Yair Goldberg, Yaron Shapira

**Affiliations:** The Department of Cardiology, Rabin Medical Center, Beilinson Hospital, Petah Tikva, Israel, Affiliated with the Gray Faculty of Medical & Health Sciences, Tel-Aviv University, Tel Aviv, Israel; The Faculty of Data and Decision Sciences, Technion – Israel Institute of Technology, Haifa, Israel

## Abstract

**BACKGROUND:** Guidelines recommend aortic valve replacement (AVR) in patients with severe aortic regurgitation (AR) based on progressive changes in left ventricular (LV) function or size. We aimed to reassess the clinical relevance of current guideline recommendations pertaining to traditional echocardiographic measurements in routine practice.

**METHODS:** Retrospective analysis of patients with severe AR who underwent serial echocardiographic follow-up over at least 18 months. The composite outcome was symptom-driven AVR, acute heart failure hospitalization, or death. We used a joint modelling approach to handle within-subject correlation and censoring.

**RESULTS:** The cohort consisted of 140 patients, with a median follow-up of 93 months (interquartile range 58–130). LV end-systolic (LVESD) and fractional shortening (FS) showed a small but statistically significant longitudinal trend, while LVEDD did not. Changes in all three parameters in parallel joint models adjusted for age and gender were consistently associated with increased risk of the composite event. Each 1 mm increase in LVESD and LVEDD was associated with a 6% and 5% increase in risk, respectively; each 1% decrease in FS corresponded to a 12% increase in risk. Only 8 (5.7%) of patients were predicted to exceed the guideline-recommended LVEDD threshold of 65 mm over 10 years. Age at onset was also a significant risk factor, with each decade increasing risk by 65% for each of the three parallel joint models.

**CONCLUSIONS:** LV parameters show modest changes over time, despite holding strong prognostic value in patients with severe AR. LVEDD, while associated with overall risk, does not predictably or significantly dilate over time in most patients. AVR decisions should be based on comprehensive clinical and volumetric assessment rather than waiting for simple linear progression to guideline cutoffs.

## INTRODUCTION

Chronic severe aortic regurgitation (AR) leads to left ventricle (LV) volume overload, resulting in progressive eccentric hypertrophy and dilatation to counter high LV wall stress^1^. Rcommendations for aortic valve replacement (AVR) related to the severity of LV dilation in AR are based on transthoracic echocardiography (TTE) parasternal long-axis linear measurement of LV at base and LV ejection fraction (LVEF)^2,3^. A LVEF less than 55% (per ACC/AHA)^2^ or less than 50% (per ESC/EACTS)^3^ carries a strong (Class I) indication for AVR. An LV end systolic diameter (LVESD) greater than 50 mm, or an indexed LVESD greater than 25 mm/m^2^, is also considered a Class I indication for AVR per ESC/EACTS guidelines, or Class IIa per ACC/AHA guidelines, and is well supported by previous studies^4–8^.

Current valve guidelines suggest that AVR may be considered in patients with severe AR based on progressive changes in left ventricular function or size, including: (1) a progressive decline in LVEF on at least three serial studies, reaching the low–normal range (55-60%)^2^; and (2) a progressive increase in LV end-diastolic diameter (LVEDD) above 65 mm^2,3^. However, to our knowledge, only two old studies by a single research group have actually looked into how these parameters change over time in patients with severe AR, and whether such changes are associated with adverse outcomes.. A 1980 study by Henry et al. followed 37 patients with repeated echocardiographic exams over a mean of 34 months^9^. A later study, published in 1991 by Bonow et al., analyzed serial echocardiographic and radionuclide angiographic studies over a mean of 8 years in 104 patients with severe AR and normal LVEF^8^. In the 1991 study, the rate of LVEDD progression did not differ between patients who remained stable and those who experienced adverse outcomes – specifically, death, heart failure symptoms, or deterioration in LV systolic function (0.3±0.8 vs. 0.4±0.8 mm/year, respectively). However, LVESD increased more in the event group (1.1 ± 1.3 vs. 0.2 ± 0.7 mm/year, p <0.005). ^8^ In the earlier study, the average LVESD change was 2.6 ± 1.2 mm/year.^9^

Given the gaps outlined above, we aimed to reassess the clinical relevance of current guideline recommendations pertaining to changes in standard echocardiographic measures. Specifically, we examined how LVEDD, LVESD, and fractional shortening (FS) change over time in patients with pure chronic moderate to severe AR or greater. Moreover, we assessed whether changes in these measures are associated with adverse clinical outcomes.

## METHODS

### Study Design and Population

We identified all consecutive patients with moderate-to-severe or severe AR on baseline transthoracic echocardiography (TTE) and at least three follow-up TTEs over more than 18 months at a large tertiary medical center between April 1994 and April 2024. Exclusion criteria were concomitant greater than mild to moderate aortic stenosis or mitral valve disease, aortic dissection, prior valve surgery, hypertrophic cardiomyopathy, complex congenital heart disease, or LV assist devices (Figure 1). The primary composite outcome included symptom-driven AVR, acute heart failure hospitalization, or all-cause mortality.

**Figure 1.**
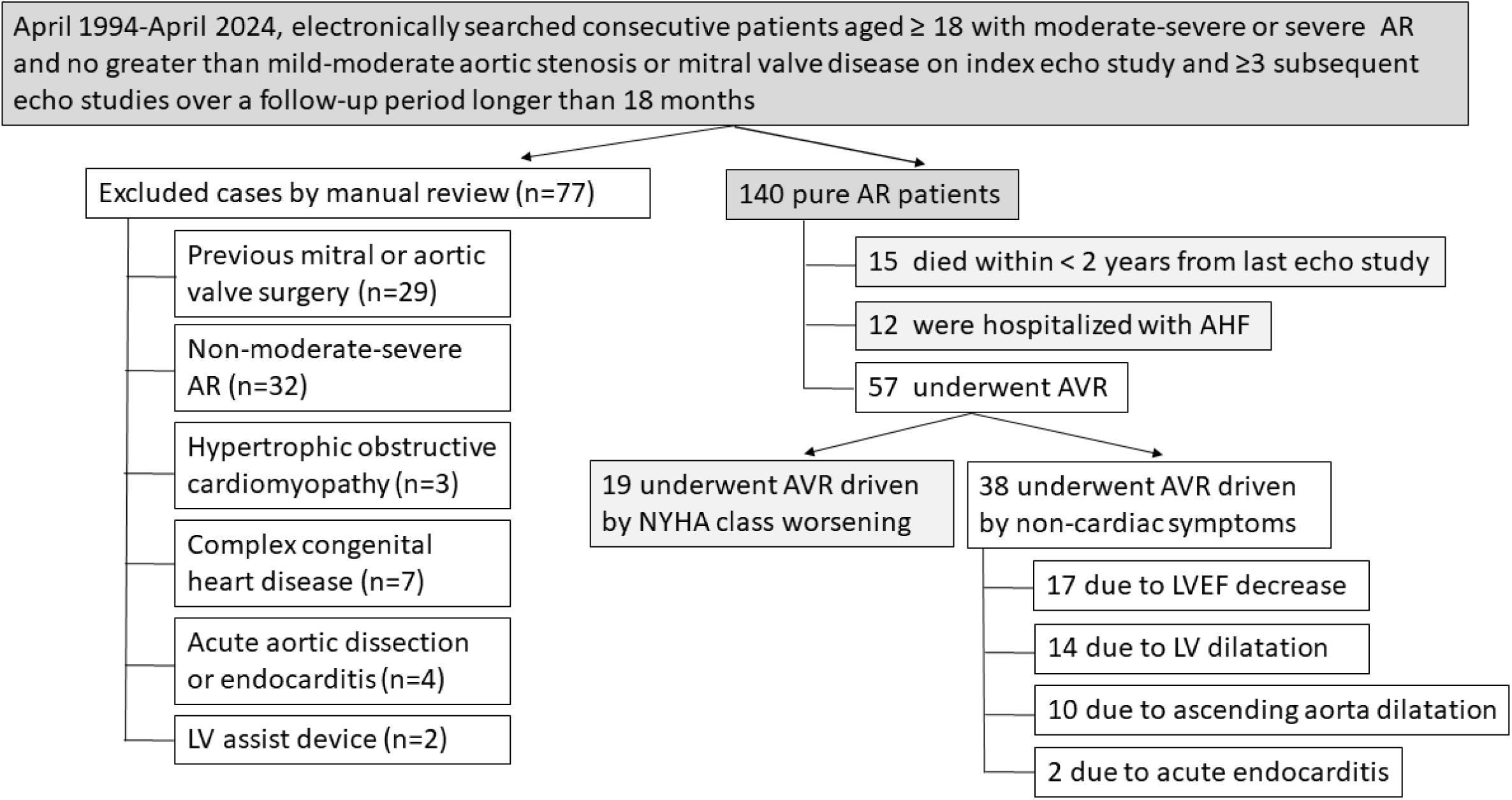
**Study flow**

We extracted baseline characteristics and echocardiographic measurements for all patients from electronic medical records and echo reports, and the causes for AVR from the medical visit records. The study was approved by the hospital’s Institutional Review Board.

### Echocardiography Studies

All patients underwent comprehensive echocardiograms using commercial instruments. All TTE studies were performed according to the guidelines of the American^2,10,11^ and European cardiology societies^3,12^. Morphology of AV (trileaflet vs. other) was documented. The inner LV cavity diameter was measured at end-diastole and end-systole at the traditional basal ventricular level on parasternal long axis view. Fractional shortening (FS) was calculated by the formula (LVEDD-LVESD)*100/LVEDD.

### Outcomes

Patients were followed for the earliest occurrence of any component of the primary composite endpoint, which included all-cause death, acute heart failure (AHF) hospitalization, or worsening heart failure (HF) symptoms prompting AVR. Worsening symptoms were defined as a deterioration of at least one New York Heart Association (NYHA) functional class despite medical management, prompting AVR, either surgical or transcatheter replacement. The definition of AHF hospitalization was standardized according to societal guidelines, following a comprehensive examination and review of electronic medical records for clinical events.

Patients who did not experience any of these events were censored at the end of follow-up or at the time of AVR if it was performed for reasons other than the appearance of cardiac symptoms. Such reasons included a decrease in LVEF, an increase in LVESD or indexed LVESD, MRI-determined LV enlargement, or dilatation of the ascending aorta or aortic root beyond accepted thresholds at the time of their referral to surgical treatment.

### Statistical Analysis

We applied an analysis that combines the following two key aspects: (1) characterizing how each patient’s echocardiographic measures, specifically FS, LVESD, and LVEDD, change over time; and (2) assessing how these measures are related to the risk of experiencing a composite event during follow-up. By fitting these two aspects together, the analysis can reveal whether specific patterns in these measures provide an early indication of future risk. We used a *joint modeling approach* implemented in the JMbayes2 package in R^13–15^, which links a linear mixed-effects model for the repeated echocardiographic measurements with a Cox proportional hazards model for time-to-event data. This framework accounts for patient-specific trajectories of echocardiographic measurements (random effects) over time and evaluates how the current value of these measurements is associated with the risk of an event. We constructed parallel joint models for FS, LVEDD and LVESD to test the robustness of our findings, adjusting for gender and age at onset.

## RESULTS

### Baseline Characteristics

We included a cohort of 140 patients meeting the inclusion criteria on initial echo study after exclusion of 77 patients (Figure 1). The distribution of the included repeated TTE studies and follow-up time is shown in Figure 2. The cohort had a median follow-up of 93 months (IQR 58–130) and a median of 9 TTE studies (IQR 6–13.5).

**Figure 2.**
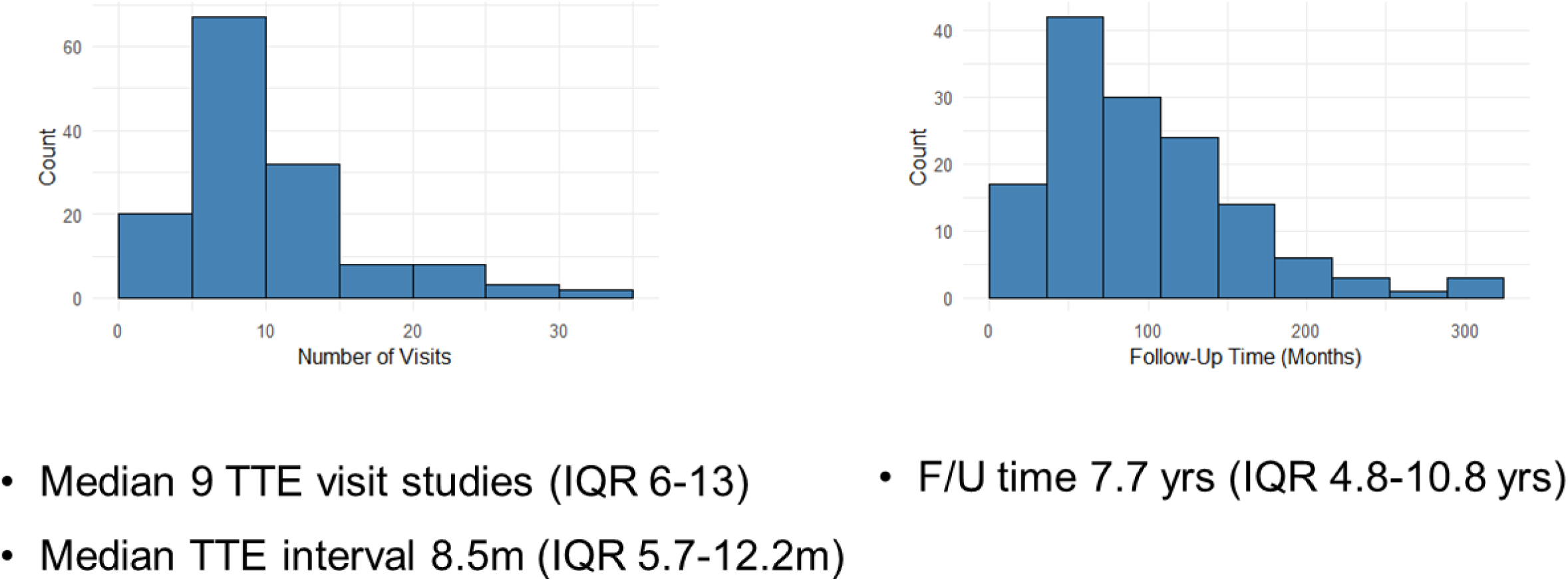
Echo studies meeting inclusion criteria distribution. Left panel: TTE count distribution across the study cohort. Right panel: follow-up time distribution across the study cohort (in months).

Baseline characteristics are presented in Table 1. Aortic valve morphology was tricuspid in 49%, bicuspid in 28%, and other/unknown in the remainder. The median of age at onset was 56 years (IQR 40–70) and 83% were male. During follow-up, 48 (34%) of patients experienced the composite outcome.

**Table 1:** Baseline data. Approximately normal variables are expressed as means ± standard deviations. Non-gaussian continuous varables are expressed as median [interquartile range]. Categorical variables are expressed as counts (percentages).

| Variable | Overall (n = 140) | Censored (n = 94) | Event (n = 46) |
| --- | --- | --- | --- |
| Follow Up in Months | 93 [58, 132] | 95 [62, 150] | 92 [57, 126] |
| Number of Echo Studies | 9.5 [6.0, 14.0] | 10.0 [7.0, 15.0] | 9.0 [6.0, 12.0] |
| Age at Baseline (Years) | 56 [40, 70] | 52 [37, 66] | 70 [51, 76] |
| Gender (Male) | 116 (83%) | 79 (84%) | 37 (80%) |
| Body Surface Area (m<sup>2</sup>) | 1.91 ± 0.20 | 1.93 ± 0.21 | 1.87 ± 0.19 |
| **Echocardiographic Measurements at Baseline** |  |  |  |
| AV Type |  |  |  |
| Tricuspid | 68 (48.6%) | 41 (43.6%) | 27 (58.7%) |
| Bicuspid | 39 (27.8%) | 34 (36.2%) | 5 (10.9%) |
| Unicuspid | 4 (2.9%) | 2 (2.1%) | 2 (4.3%) |
| Quadricuspid | 1 (0.7%) | 1 (1.1%) | 0 (0%) |
| Unknown | 28 (20%) | 16 (17%) | 12 (26.1%) |
| Ascending Aorta Diameter (cm) | 3.97 ± 0.67 | 3.92 ± 0.69 | 4.13 ± 0.57 |
| LVEDD (mm) | 57 ± 7 | 56 ± 6 | 58 ± 8 |
| LVEDS (mm) | 37 ± 7 | 37 ± 6 | 38 ± 9 |
| Interventricular Septum (cm) | 1.12 ± 0.22 | 1.11 ± 0.20 | 1.15 ± 0.25 |
| Posterior Wall (cm) | 1.04 ± 0.18 | 1.03 ± 0.18 | 1.05 ± 0.19 |
| FS (%) | 35 ± 7 | 35 ± 6 | 36 ± 9 |
| LV ejection fraction (EF) |  |  |  |
| Good (55-65%) | 116 (82.9%) | 84 (89%) | 32 (69.5%) |
| Preserved (50-54%) | 12 (8.6%) | 4 (4.3%) | 8 (17.4%) |
| Mild (45-49%) | 10 (7.1%) | 6 (6.4%) | 4 (8.7%) |
| Mild to Moderate (30-34%) | 1 (0.7%) | 0 (0%) | 1 (2.2%) |
| Unknown | 1 (0.7%) | 0 (0%) | 1 (2.2%) |
| RV systolic pressure (mm Hg) | 24 [21, 30] | 25 [21, 30] | 22 [20, 30] |
| Mitral Regurgitation Grade |  |  |  |
| none or mild | 122 (87.2%) | 80 (85.1%) | 42 (91%) |
| mild to moderate | 17 (12.1%) | 13 (13.8%) | 4 (8.7%) |
| Unknown | 1 (0.7%) | 1 (1.1%) | 0 (0%) |
| Aortic Stenosis Grade |  |  |  |
| none or mild | 138 (99%) | 93 (99%) | 45 (98%) |
| mild to moderate | 2 (1.4%) | 1 (1.1%) | 1 (2.2%) |
| Tricuspid Regurgitation Grade |  |  |  |
| none or mild | 131 (93.6%) | 87 (92.5%) | 44 (95.6%) |
| mild to moderate | 7 (5%) | 6 (6.4%) | 1 (2.2%) |
| Unknown | 2 (1.4%) | 1 (1.1%) | 1 (2.2%) |

| **Variable** | **Overall (n = 140)** | **Censored (n = 94)** | **Event (n = 46)** |
| --- | --- | --- | --- |
| **Outcomes** |  |  |  |
| Censored | 94 (67%) | 94 (100%) | 0 (0%) |
| Acute Heart Failure | 12 (8.6%) | 0 (0%) | 12 (26%) |
| Aortic Valve Replacement | 19 (14%) | 0 (0%) | 19 (41%) |
| Death | 15 (11%) | 0 (0%) | 15 (33%) |

Forty-eight patients (34%) experienced the composite event: 15 deaths, 19 symptom-driven AVRs, and 12 HF hospitalizations. Thirty-eight additional AVRs occurred for non-symptomatic indications (e.g., LV dilation, LVEF decline, aortic enlargement) and were therefore censored. The AVR causes in these censored patients are detailed in Figure 1.

Figure 3 illustrates FS measurements over time for four example patients, each plotted alongside their best-fitting linear trend line. In Figures 3A–3C, a vertical dashed line marks the timing of the clinical event. In Figure 3D, the patient did not experience any event, and follow-up was censored.

**Figure 3.**
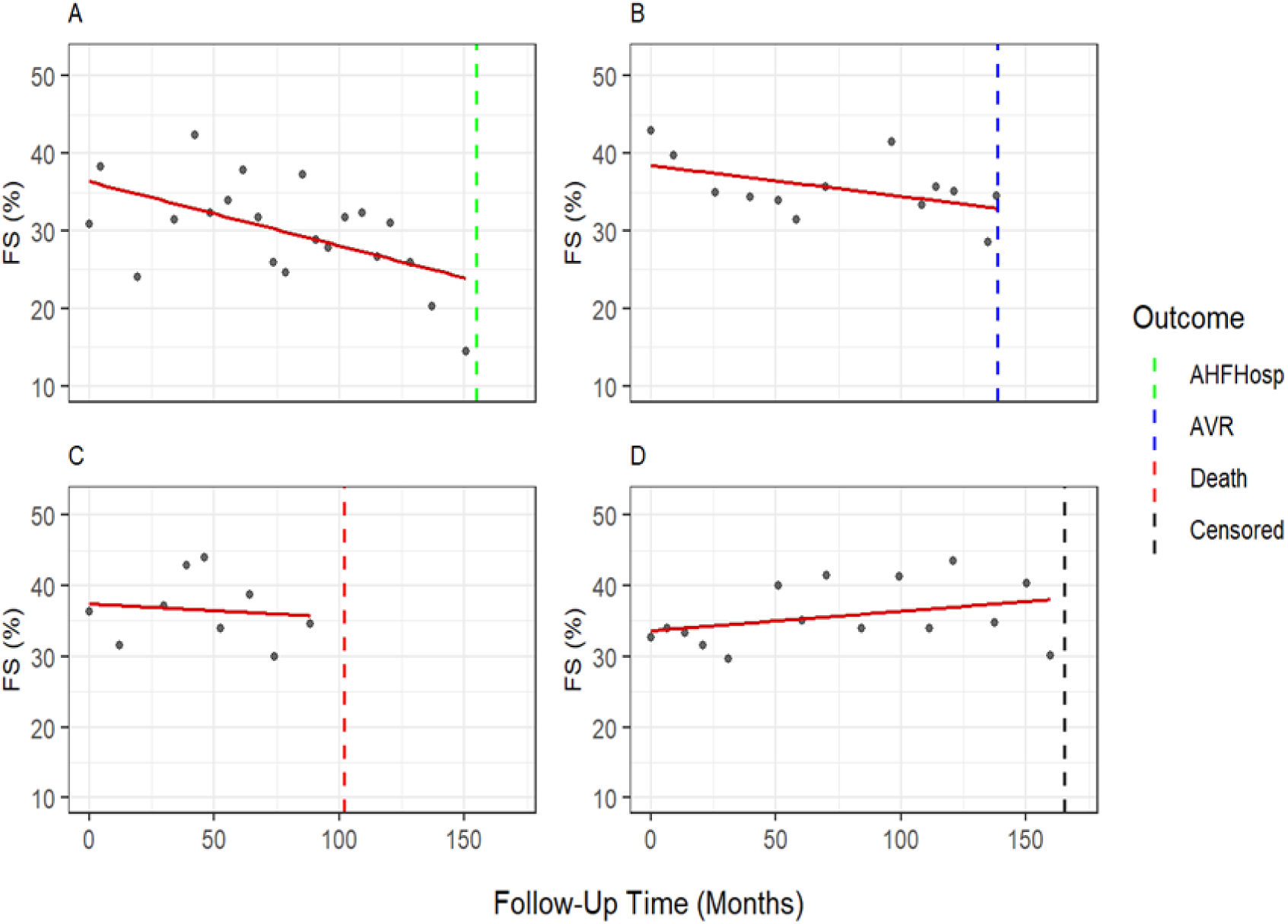
Individual FS Measurements with linear fit and event time indication in four illustrative patients. The linear fit is shown for reference only and was not used in the statistical analysis. See text for further explanations.

### Joint model using fractional shortening (FS) as the longitudinal variable

Table 2 summarizes the joint model results with FS as the main echocardiographic measure. The longitudinal model accounts for repeated measurements from each patient and estimates how FS is affected by time, age, and gender. In this analysis, a linear trend is fitted to each individual’s FS values over time, and Table 2.I shows the average of these trends across all patients.

**Table 2:** Joint Model for Fractional Shortening (FS)

| **Variable** | **Mean** | **StDev** | **2.5%** | **97.5%** | **P** |
| --- | --- | --- | --- | --- | --- |
| **I. Longitudinal Submodel Results** |  |  |  |  |  |
| **Intercept (%)** | 38.15 | 1.58 | 35.07 | 41.31 | <0.01 |
| **Follow-Up in months** | -0.02 | 0.00 | -0.03 | -0.01 | <0.01 |
| **Age at onset (Years)** | -0.03 | 0.02 | -0.08 | 0.01 | 0.17 |
| **Gender (Male)** | -1.63 | 1.07 | -3.72 | 0.49 | 0.13 |
| **Sigma** | 5.10 | 0.11 | 4.89 | 5.31 | <0.01 |
| **II. Survival Submodel Results** |  |  |  |  |  |
| **FS** | 0.05 | 0.01 | 0.02 | 0.08 | <0.01 |
| **Age at Onset in years** | -0.49 | 0.40 | -1.25 | 0.36 | 0.23 |
| **Gender (Male)** | -0.11 | 0.04 | -0.19 | -0.04 | <0.01 |

The average FS at baseline TTE was around 38%. Over time, FS tended to decline significantly but slightly, by about 0.02% per month. Older age at baseline TTE was linked to slightly lower FS, and men had FS values about 1.62% lower than women on average; however, neither of these differences was statistically significant. The variability of individual FS measurements around their expected trends, represented by the sigma value, was approximately 5%. Overall, FS declined modestly over time across the population; however individual patterns varied widely, and no clear differences were observed by age or gender.

Figure 4 presents subject-specific predicted FS trajectories (in blue) for the four illustrative individuals shown in Figure 3, alongside the overall population trend (in black), as estimated by the linear mixed-effects model. The shaded areas represent a 95% confidence interval (±1.96 standard deviations) around each individual’s predicted trajectory. As shown, measurement variability introduces noticeable noise into the estimated individual trajectories. The model pulls these individual trends toward the population average, which stabilizes the predictions but results in wider confidence intervals, reflecting greater uncertainty.

**Figure 4.**
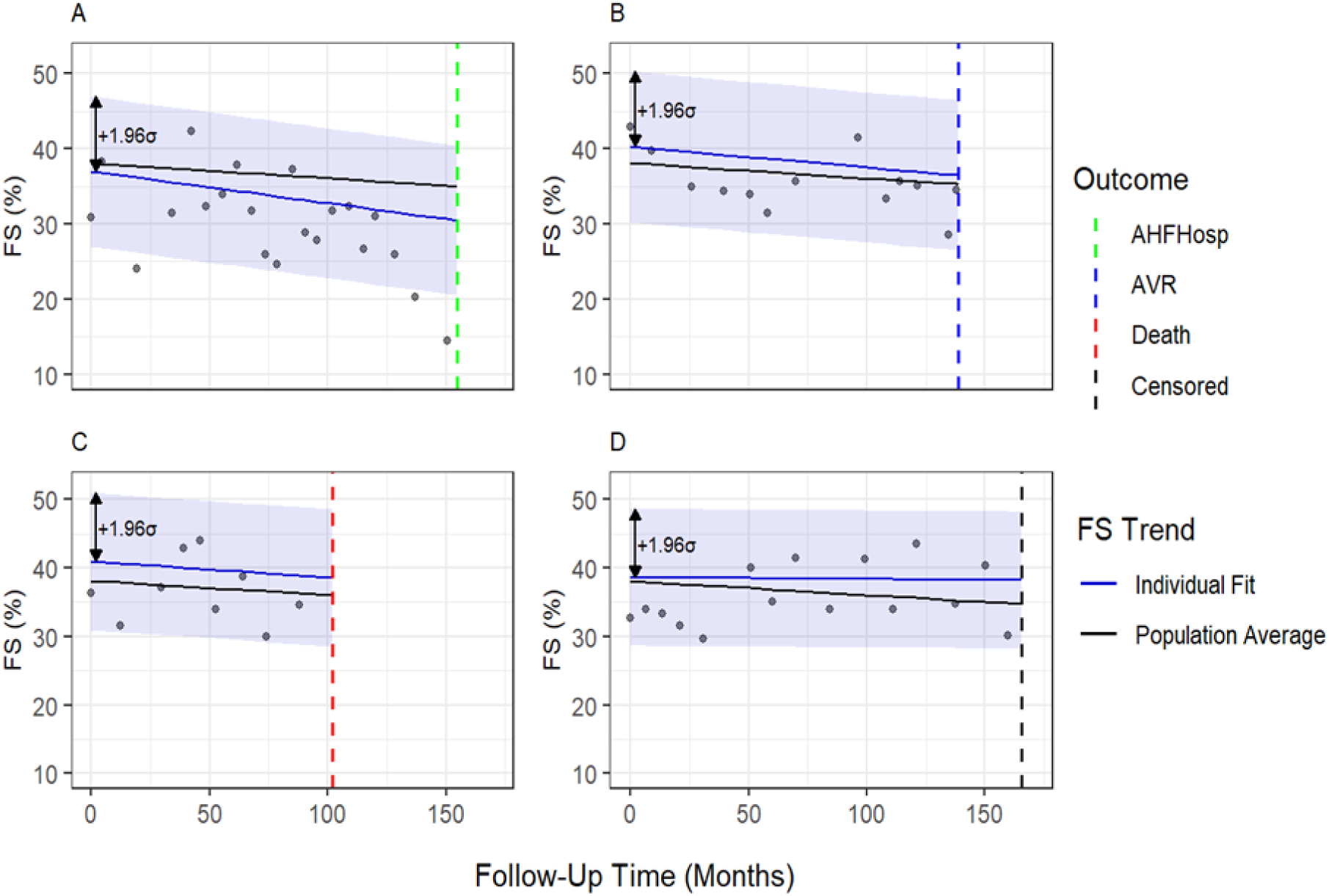
Comparison of population-level trend and individual-specific trajectories of FS for the four patients in prior figure (shown in blue) alongside the overall population trend (in black), as estimated by the linear mixed-effects model. The shaded areas represent a 95% confidence interval (±1.96 standard deviations) around each individual’s predicted trajectory. See text for further explanations.

The survival submodel in Table 2.II estimates how patient age, gender, and current FS level (predicted from the longitudinal submodel) are associated with the hazard of experiencing the composite event. The coefficients presented correspond to effects on the log-hazard scale. Exponentiation of these coefficients yields hazard ratio (HR) estimates. The results show that lower FS values are significantly associated with increased event risk.

Specifically, the coefficient of –0.11 for FS implies a HR of approximately 1.12, indicating a 12% higher risk of experiencing the composite outcome per 1% decrease in FS. A 4% decrease in FS (approximately 10% from baseline average value) corresponded to an HR of 1.55. Baseline age also showed a statistically significant effect, with HRs of 1.05 per year (5% higher risk), 1.28 per five years, and 1.65 per ten years. Gender was not significantly associated with the risk of experiencing the composite event.

### Joint model using LVESD as the longitudinal variable

Table 3 summarizes the joint model results with LVESD as the main echocardiographic measure. The average LVESD at baseline TTE was approximately 35 mm. Over time, LVESD showed a small but statistically significant increase of 0.01mm/month, suggesting a gradual widening of the left ventricle during follow-up. Age at onset was not meaningfully associated with LVESD levels. However, male patients had significantly higher LVESD values than females, by an average of 3.2 mm. The typical variability in individual LVESD measurements around the expected trend, represented by the sigma value, was estimated at 3.37 mm.

**Table 3:**
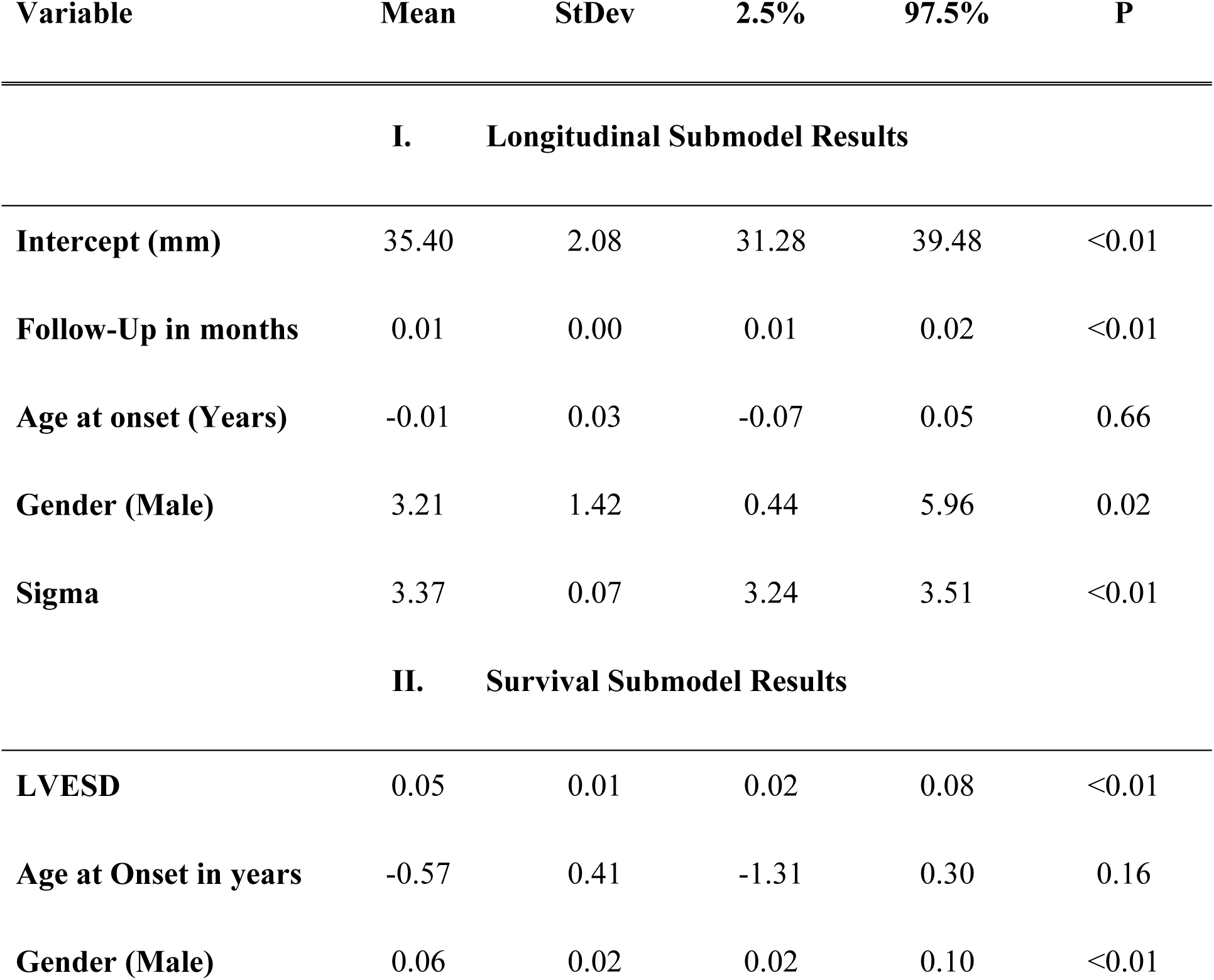
Joint Model for LVESD.

The survival submodel in Table 3.II shows that patients with larger LVESD values were at higher risk of experiencing the composite outcome, with a HR of 1.06 per 1 mm increase in LVESD (6% higher risk) and 1.20 per 3 mm increase (i.e. an increase close to 10% from average baseline value).

Baseline age was also a significant factor, with an identical effect to that in the FS-based parallel analysis. As with the FS analysis, there was no meaningful difference in risk between male and female patients.

### Joint model using LVEDD as the longitudinal variable

Table 4 summarizes the LVEDD joint model. The results indicate that, on average, LVEDD remained stable over time, as the effect of follow-up duration was close to zero and not statistically significant. Thus, the average LVEDD value of approximately 56 mm, indicated by intercept at Table 4, reflects the overall population mean across the entire follow-up period and not just at baseline. LVEDD was not meaningfully associated with age at baseline TTE. Male patients had significantly higher LVEDD values than females, by about 4.2 mm on average. The typical variability in individual LVEDD measurements around the expected trajectory of each patient, represented by the sigma value, was estimated at 3.38 mm.

**Table 4:** Joint Model for LVEDD.

| Variable | Mean | StDev | 2.5% | 97.5% | P |
| --- | --- | --- | --- | --- | --- |
| **I. Longitudinal Submodel Results** |  |  |  |  |  |
| **Intercept (mm)** | 55.99 | 1.97 | 52.19 | 59.97 | <0.01 |
| **Follow-Up in months** | -0.01 | 0.00 | -0.01 | 0.00 | 0.14 |
| **Age at onset (Years)** | -0.04 | 0.03 | -0.10 | 0.02 | 0.18 |
| **Gender (Male)** | 4.19 | 1.35 | 1.53 | 6.84 | <0.01 |
| **Sigma** | 3.38 | 0.07 | 3.24 | 3.51 | <0.01 |
| **II. Survival Submodel Results** |  |  |  |  |  |
| **LVEDD** | 0.05 | 0.01 | 0.03 | 0.08 | <0.01 |
| **Age at Onset in years** | -0.56 | 0.40 | -1.32 | 0.28 | 0.18 |
| **Gender (Male)** | 0.05 | 0.02 | 0.01 | 0.10 | 0.02 |

The findings of the survival submodel in Table 4.II closely mirror those from the LVESD model: larger LVEDD values were associated with a higher risk of experiencing the composite outcome, with an HR of 1.05 per 1 mm increase in LVEDD (5% higher risk) and 1.16 per 3 mm increase (16% higher risk). Age was again a significant predictor, with an identical effect to that observed in both LVESD- and FS-based models. Consistent with those models, no meaningful difference in risk was observed between male and female patients.

## DISCUSSION

In our study, we revisited the value of scrutinizing repeated simple 2D echocardiographic measurements, as advocated by the American and European guidelines. We evaluated this in a modern real-world scenario of long-term clinical follow-up of patients with significant AR, using a large retrospective database from a tertiary medical center. Substantial noise was introduced by technical variability, as TTE studies were performed by different cardiac sonographers on different commercial echo machines. In addition, patients underwent varying numbers and frequencies of TTE studies over the follow-up period, further contributing to heterogeneity. Nevertheless, this variability also allowed us to test the robustness of widely used real-world practice tools in a representative clinical setting. By constructing parallel statistical joint models for FS, LVESD and LVEDD, we were able to evaluate the association with outcomes in this heterogeneous setting. The longitudinal results of all the models were consistent with the overall variability observed in the repeated measurements, as reflected by the large variability estimates around each patient’s assessed trajectory.

All three echocardiographic measurements (LVESD, FS, and LVEDD) were significantly associated with an increased risk of the composite adverse outcome. A modest but consistent population-level longitudinal trend was observed for LVESD (increase) and FS (decline), whereas LVEDD showed no consistent trend over time. The observed LVESD increase rate was similar to that reported by Bonow et al. in AR patients who developed death, symptoms, or left ventricular dysfunction^8^.

Based on the models intercepts, male patients had, on average, larger hearts than female patients (about 3 mm greater LVESD and about 4 mm greater LVEDD), consistent with known physiological gender differences^16^. However, gender was not associated with differences in the composite risk in any of the joint models.

Baseline age was a significant predictor for the composite event in all joint models, with a consistent hazard ratio of 1.05 per additional year, or 1.28 per five additional years. Despite a higher risk of post-operative mortality and complications in elderly patients^17^, this finding supports AVR consideration in older patients who meet guideline-based indications (i.e., LVESD increase or LVEF decline) and lack significant comorbidities.

Although TTE basal linear LV diameters are useful for prognosticating patients with chronic AR, they may come at the risk of poorly capturing the true LV remodeling and eccentric LV dilation that commonly occurs^18,19^. The LVESD is a unidimensional estimate of LV end-systolic volume, and LVEDD an estimate of LV end-diastolic volume, with only moderate correlation between the linear dimensions and volumes in AR patients^19–21^. The reason that in our study we found that LVESD increases modestly but significantly over a long follow-up period whereas LVEDD did not may be related to their different determinants. The LV end-diastolic volume only captures the effect of preload whereas LV end-systolic volume reflects both the effect of preload and the systolic performance and is therefore a better parameter to assess LV remodeling in AR patients^22,23^. In fact, several studies showed that LV end-systolic volume (or indexed volume) predicts postoperative outcomes, survival, and symptom progression better than LV end-diastolic volume in AR^19,20,24^. It has been shown that linear dimensions measured at the midventricular level, rather than the traditional basal level, are more closely correlated with volumetric measurements of LV size, and LVEF^18^, but midventricular dimensions were not recorded systematically in our study.

Based on our results, we evaluated the clinical relevance of the current guideline recommendation to perform AVR when LVEDD exceeds 65 mm^2,3^. Using the fitted longitudinal model, we predicted individual LVEDD trajectories over a 10-year follow-up period. which reflects a reasonable extension of the observed median follow-up of 7.75 years. Model-based predictions indicated that only 8 out of 140 patients (5.7%) were expected to exceed the 65 mm threshold at any point during this period. Of these, four patients had persistently elevated LVEDD values above 65 mm, while three patients started with values above 65 mm but showed decreasing trajectories over time. Importantly, only one patient crossed the 65 mm threshold during follow-up, representing upward progression. These findings question the applicability of a fixed LVEDD cutoff as a standalone intervention criterion. Similarly, in a recent study of 365 patients with severe AR who underwent AVR, de Meester et al. found no difference in post-AVR survival between those with LVEDD below versus above 65 mm^7^. However, their design and outcome measure (post-operative mortality) differed from ours (composite endpoint pre-AVR), limiting direct comparability.

FS became a popular measure of LV function with the advent of M-mode echocardiography in the late 70’s^25,26^ . In a small old study of patients with AR, FS was found to decrease to a lesser extent in asymptomatic patients compared with those who developed dyspnea^27^. FS has well known limitations, as it is a linear parameter which may not accurately represent overall cardiac function in patients with regional wall motion abnormalities or abnormal ventricular activation^16^. However, it correlates with LVEF and, in this study, served as a surrogate for LVEF in AR patients since precise quantitative LVEF was unavailable in most TTE studies in our database.

The decline in FS over time observed in our patients, and increased adverse risk associated with FS decrease resonate with the known LVEF decline and its prognostic value in AR patients in literature^4,8,28,29^. These findings also align with a 1980 study showing that preoperative FS < 25% identified patients at risk of post-AVR death related to congestive heart failure^9^. Interestingly, although 12 patients (8.6%) in our study had reduced LVEF, the median FS on baseline TTE was 35%. This value is within normal range according to old data for healthy subjects of similar age^25,26^, and is higher than the average FS of 30% reported in a prior study of 50 patients with AR^9^.

### Limitations

A major limitation of our study is its long-term retrospective nature, which is inherently associated with variability in the characteristics of TTEs performed related to clinical and sonographer factors. The joint modeling approach we utilized in this setting enabled us to meet the aforementioned challenges.

Although the dataset contained many potential variables, the relatively small sample size (140 patients) limited the complexity of the statistical models. Including more variables would have made them prone to overfitting and unstable results. Therefore, the models included only a few covariates and did not jointly model multiple time-dependent variables.

Important parameters for AR assessment and management, including MRI-derived volumes and LVEF, as well as echo-derived biplane method-of-disks volumes and LVEF and midventricular diameters (which, as mentioned earlier, correlate better with volumetric measurements of LV size), were unavailable for many patients in our cohort. In most patients in our database, only qualitative (categorical) LVEF was recorded. Therefore, we used FS as a continuous, though less precise, surrogate measure of LV systolic function instead of quantitative LVEF.

## CONCLUSIONS

In this real-world cohort of patients with moderate-to-severe or severe AR, LVESD increased modestly but significantly during long-term follow-up, whereas LVEDD showed no consistent population-level change. Nevertheless, higher LVESD and LVEDD, as well as an older age, were associated with increased risk of the composite outcome. FS, used as a surrogate for LVEF, declined modestly but significantly over time, and lower FS values were also associated with higher risk. Although men had larger average LVESD and LVEDD values than women, gender was not associated with adverse composite outcomes.

Taken together, our findings show that LVEDD, LVESD, and FS are significant predictors of adverse outcomes, supporting their prognostic role in the longitudinal assessment of patients with AR. However, the relatively small effect sizes and considerable variability in individual trajectories indicate that AVR decisions should be based on a comprehensive clinical assessment rather than changes in these parameters alone. The model predicted that only a small proportion of patients would surpass the 65 mm LVEDD threshold during follow-up, and in most of these cases this did not reflect a progressive increase over time. These findings suggest that this criterion may have limited utility as a sole trigger for intervention in routine follow-up.

## CLINICAL PERSPECTIVE

### What Is New?

Linear measurements of left ventricle diameters in AR is simple and has been widely used and recommended in clinical practice, but without robust evidence as to consistent progressive changes over time in the individual patient. Our study confirms that LVESD and fractional shortening demonstrate modest longitudinal changes and strong prognostic associations in patients with severe AR, whereas LVEDD, while associated with risk, does not change significantly over time.

### What Are the Clinical Implications ?

The relatively small effect sizes and considerable variability in individual trajectories underscores the importance of basing AVR decisions on a comprehensive clinical assessment complemented by volumetric measures that have been well-established in recent studies rather than relying on changes in traditional linear LV parameters.

## Data Availability

All data referred to in the manuscript is readily available upon request.

---

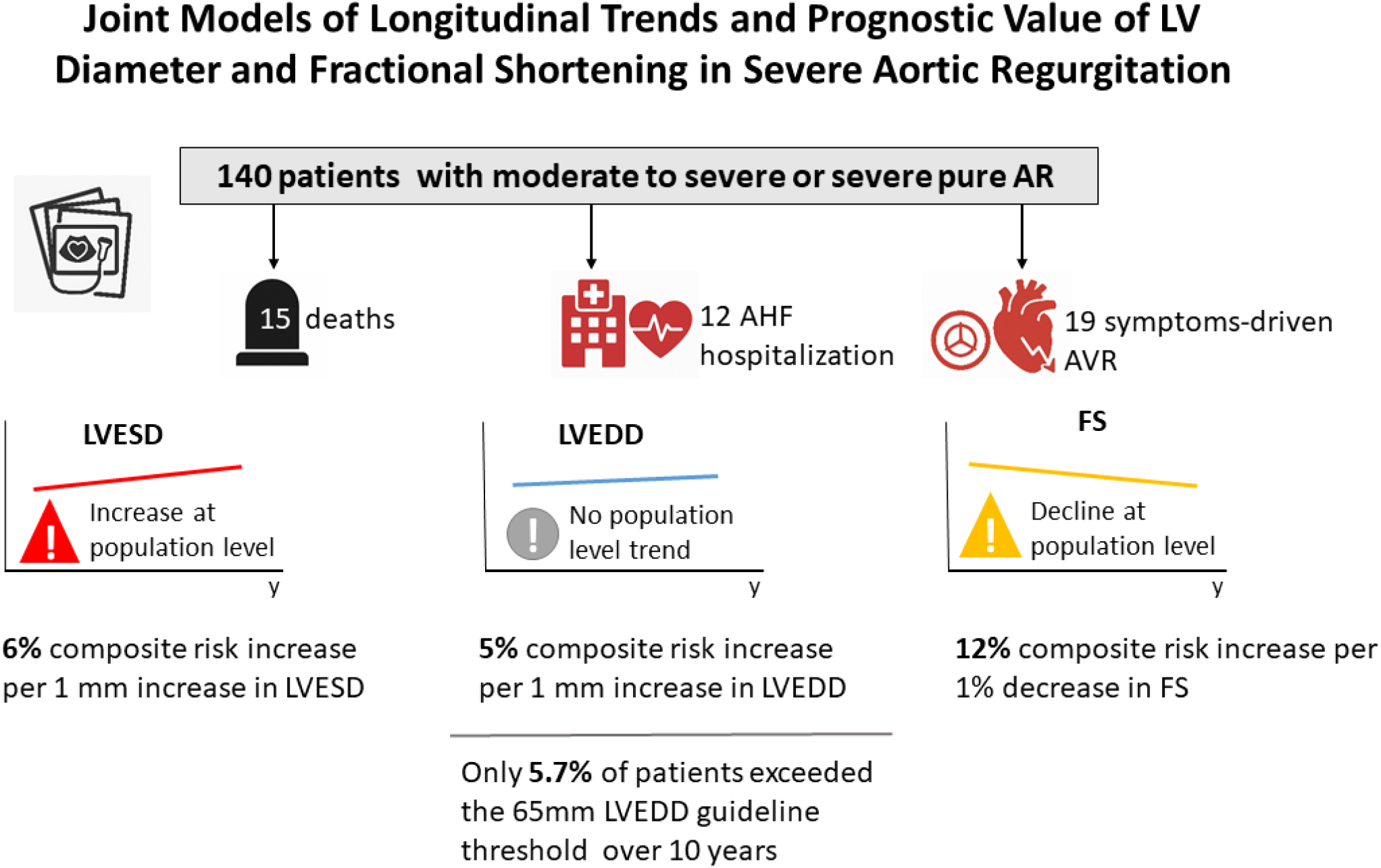

